# Multi-omics endotype of preterm infants with bronchopulmonary dysplasia and pulmonary hypertension

**DOI:** 10.1101/2022.11.03.22281890

**Authors:** Roopa Siddaiah, Christiana Oji-Mmuo, Vincent Aluquin, Yuka Imamura Kawasawa, Ann Donnelly, Dustin Rousselle, Nathalie Fuentes, Eric D. Austin, Patricia Silveyra

## Abstract

**Rationale:** Pulmonary hypertension associated with bronchopulmonary dysplasia is a severe complication of preterm birth resulting in high mortality of up to 50% within the first 2 years of life. There is a direct relationship between bronchopulmonary dysplasia severity and incidence of associated pulmonary hypertension. However, it is challenging to clinically characterize severe bronchopulmonary dysplasia with and without pulmonary hypertension and there is need for better understanding of the two entities.

**Objectives:** To identify markers to help understand biological processes and endotype characterization of infants with pulmonary hypertension associated with bronchopulmonary dysplasia in tracheal aspirates.

**Methods:** We conducted multi-omic analysis of tracheal aspirates via miRNA PCR arrays, RNA sequencing and mass spectrometry proteomics in preterm infants with severe bronchopulmonary dysplasia with (n=21) and without (n=25) pulmonary hypertension.

**Results:** Our study analysis revealed 12 miRNAs (hsa-miR-29a, has-miR-542-3p, has-miR-624, has-miR-183, hsa-miR-501-3p, hsa-miR-101, hsa-miR-3131, hsa-miR-3683, hsa-miR-3193, hsa-miR-3672, hsa-miR-3128, and hsa-miR-1287); 6 transcripts (IL6, RPL35P5, HSD3B7, RNA5SP215, OR2A1-AS1, and RNVU1-19), and 5 proteins (CAPS, AAT, KRT5, SFTPB, and LGALS3BP) with significant differential expression in preterm infants with severe lung disease with pulmonary hypertension when compared to infants with severe lung disease but no pulmonary hypertension. Pathway analysis of the integrated multi-omic expression signatures revealed NFkB, VEGF, SERPINA1, IL6 and ERK12 as target molecules for miRNAs, and angiogenesis and hyperoxia stress as recurrent pathways of individual markers.

**Conclusion:** Our multi-omic analysis of tracheal aspirates revealed a comprehensive thumbprint of miRNAs, mRNAs and proteins that could help endotype infants with severe lung disease and pulmonary hypertension.

## Introduction

With recent advances in neonatal care, there is improved survival of extremely premature babies with very low birth weight, although significant reductions in complications of bronchopulmonary dysplasia (BPD) remain lacking (1). The pathogenesis of BPD is multifactorial and clinical phenotypes highly variable. While halted alveolar and pulmonary vascular growth have been identified as key players in the pathophysiology of “new BPD”, one of its more severe complications is pulmonary hypertension associated with BPD (BPD-PH) which carries significant high morbidity and mortality (2).

The true incidence of BPD-PH in preterm babies is unknown. While the prevalence of BPD-PH is roughly one-third of all BPD cases, it is much higher in severe BPD (17%-43%) (3-5). Strictly speaking, the diagnosis of BPD is made around 36 weeks of gestational age (6, 7), while BPD-PH presents in both infancy and young childhood. Development of BPD-PH leads to increased number of days in the neonatal intensive care (NICU), increased number of days on ventilatory support, and oxygen requirement (8). In addition, a large number of these babies also require tracheostomy and home ventilatory support for prolonged periods (9). Once infants are discharged home, they continue to have high mortality and morbidity with increased hospital readmissions in the first 2 years of life (10, 11). Multiple factors including prenatal and postnatal environmental stressors such as inflammation, ventilator associated lung injury, hypoxic and hyperoxic stress, and others have been implicated in the etiology of BPD-PH (5, 12).

BPD is defined and classified based on the therapeutic need for respiratory support at 36 weeks postmenstrual age (PMA) and its outcomes (6, 7, 13). The disease phenotype of severe BPD (sBPD) is variable, hence differentiating its endotype is challenging in the absence of validated biomarkers. For example, Wu et al. recently showed that greater than 60% of infants with sBPD had pulmonary vascular disease (14). There is also significant overlap in the underlying pathophysiology of both sBPD and BPD-PH. Due to the complex array of contributing factors, the prevention, clinical diagnosis, and effective management of BPD-PH are challenging. The development of lung alveoli is dependent on pulmonary vascular development, and any disruption of vascular growth and signaling leads to reduction in alveolar development (15, 16). Studies have also indicated that postnatal imbalance between pro- and antiangiogenic factors triggered by inflammation, and oxidative and hypoxic stressors also contribute to the development of BPD-PH (17-19). While maternal preeclampsia, chorioamnionitis and small for gestational age are identified as the predominant risk factors for developing BPD-PH (17, 20) and could potentially predict its development, these are also factors that have been associated with development of sBPD. This limits their potential to be used as specific markers for BPD-PH prediction, thus novel markers are needed.

Although there have been some indications of hypoxic stress injury and vascular growth factors playing a role in the development of BPD-PH, there is currently a critical gap in our knowledge of the specific molecular pathways and mechanisms involved. In the current study, we conducted a multi-omics (proteomics, miRNAs, transcriptomics) profiling of tracheal aspirates of infants with sBPD and BPD-PH in order to identify key biomarkers resulting from convergence of multiple pathophysiological processes occurring in both conditions.

## Methods

### Subjects and samples

Once approved by the Penn State Health institutional review board (STUDY 00000482), infants born < 28 weeks of gestation were screened, and informed consent obtained from parents. We included infants with diagnosis of severe BPD type II (13) or grade 3 BPD based on 2019 NICHD/NRN classification (7). Infants with congenital anomalies, chromosomal syndromes, and cyanotic cardiac defects were excluded. BPD-PH was defined based on echocardiogram features of elevated right sided pressures including interventricular septal flattening, tricuspid regurgitation jet velocity and annular plane systolic excursion, and pulmonary artery acceleration time. Tracheal aspirate (TA) was collected from 46 infants on ventilator, by instilling 1ml of normal saline and suctioning of endotracheal secretions. Samples were immediately stored at -80°C. Relevant clinical information was obtained by chart review. Samples were grouped into sBPD (n=25) (infants on invasive mechanical ventilation at 36 weeks PMA and no signs of PH on echocardiogram) and BPD-PH (n=21) (infants on invasive mechanical ventilation at 36 weeks PMA with signs of PH on echocardiogram).

### MiRNA profiling

miRNAs were extracted and analyzed from 500 μL of TAs and analyzed with a miRNA array as described previously (21). Datasets were uploaded to GEO: https://www.ncbi.nlm.nih.gov/geo/query/acc.cgi?acc=GSE205138. The top differentially expressed miRNAs were validated in a subset of samples (n=17), with commercial miRNA assays (GeneCopoeia) and hsa-miR-16-1-3p as normalization control (22). Differential expression was calculated using the 2-ΔΔCT method (23), and significant differences determined by Student’s t-test using GraphPad Prism.

### Transcriptomics analysis

Library preparation and sequencing were performed from a sample subset, sBPD (n=15) and BPD-PH (N=7), at the Penn State Health genomics core as previously described (21). Sample counts were uploaded to GEO (https://www.ncbi.nlm.nih.gov/geo/query/acc.cgi?acc=GSE156028). The expression of the top differentially expressed gene (IL6) was validated by qPCR in a subset of samples (n=15).

### Proteomics analysis

TAs containing 10 μg of protein were pooled and iTRAQ labeled according to published protocols (24) at the Penn State Health proteomics core. Peptides were identified and quantified using the Paragon Algorithm in ProteinPilotTM 4.5beta software (ABSciex) (25). Stringent Local false discovery rate (FDR) estimation was calculated from the Proteomics System Performance Evaluation Pipeline algorithm (26). *Data analysis*: For miRNA arrays, statistical analyses were performed with the R software using the Bioconductor limma package. For transcriptomics, filtered reads were aligned to the human reference genome (GRCh38) using HISAT2 (version 2.1.0) (27), and the resulting de-duplicated reads were summarized to each gene using HTSeq (28). Differential gene expression analysis was conducted with the edgeR package on R (29), after normalizing data with the TMM method (30). Heatmaps were generated with the nonnegative matrix factorization package (31). For proteomics, data was analyzed using the Scaffold Q+ for peptide and protein identifications, and probabilities were assigned by the Protein Prophet algorithm (32). Channels were corrected in all samples according to the i-Tracker algorithm (33). Normalization was performed iteratively on intensities, as described in Statistical Analysis of Relative Labeled Mass Spectrometry Data from Complex Samples Using ANOVA (34).

### Pathway Analysis

The Ingenuity Pathway Analysis (IPA) software (QIAGEN) was used to identify miRNA, mRNA and protein interaction networks, predicted target genes, and molecular functions based on prediction scores (35). The core analysis functionality was performed for direct and indirect relationships using the Ingenuity knowledge base.

## Results

### Patient demographics

TAs from 46 preterm infants needing mechanical ventilation support were collected. Of there, 25 infants had sBPD without evidence of BPD-PH on echocardiogram and 21 infants had clinical features characterized as sBPD with evidence of BPD-PH on echocardiogram. A summary is shown in **Table 1**. No statistically significant differences were found for measured variables including gestational age (GA) at birth, birth weight, day of life at sample collection time, male sex, and racial/ethnicity group. Birth delivery by C-section was higher in the sBPD group compared to BPD-PH. Gestational age at the time of sample collection was significantly higher in the BPD-PH group **(Table 1)**.

**Table 1.** Patient demographics at study enrollment

| **Characteristic** | **sBPD**<br>**(n = 25)** | **BPD+PH**<br>**(n = 21)** | **p-Value** |
| --- | --- | --- | --- |
| **Gestational age (GA) at birth,**<br>**weeks (mean ± SD)** | 25.88 ± 1.53 | 25.41 ± 1.82 | 0.345 |
| **GA at sample collection, weeks**<br>**(mean ± SD)** | 36.15 ± 10.88 | 44.06 ± 14.73 | 0.042 |
| **Birth weight, grams (mean ±**<br>**SD)** | 747 ± 206 | 647 ± 208 | 0.112 |
| **Male sex, % (n)** | 56 (14) | 62 (13) | 0.345 |
| **Fractional inspired oxygen**<br>**during sample collection (mean**<br>**± SD)** | 0.37 ± 0.13 | 0.35 ± 0.10 | 0.572 |
| **Delivered via C-section, % (n)** | 88 (22) | 43 (9) | 0.002 |
| **Racial/ethnic group, % (n)** |  |  |  |
| **Non-Hispanic White** | 52 (13) | 57 (12) | 0.845 |
| **Non-Hispanic Black** | 4 (1) | 5 (1) | 1.0 |
| **Hispanic** | 24 (6) | 14 (3) | 0.477 |
| **Asian** | 12 (3) | 0 (0) | 0.240 |
| **More than one race** | 8 (2) | 24 (5) | 0.220 |

### miRNA analysis

A total of 1,059 miRNAs were expressed in TA samples from sBPD and BPD-PH **(Figure 1)**. A list of miRNAs is shown in **Supplementary Table 1**, and a volcano plot is shown in **Figure 2**. After accounting for false discovery rate (FDR) < 0.05, the PCR array analysis revealed differential expression of 12 miRNAs with |FC| > 2 between the BPD-PH and sBPD samples (**Table 2**). Of these, 9 miRNAs (hsa-miR-29a, hsa-miR-542-3p, hsa-miR-624*, hsa-miR-3193, hsa-miR-3672, hsa-miR-3683, hsa-miR-501-3p, hsa-miR-101*, and hsa-miR-3128) had significantly higher expression in the BPD-PH group, compared to the sBPD group, whereas 3 miRNAs (hsa-miR-183*, hsa-miR-3131, and hsa-miR-1287) had significantly lower expression in BPD-PH vs. sBPD (**Figure 3**). Validation qPCR experiments in a select subset of samples confirmed upregulation of hsa-miR-29a, hsa-miR-542-3p, hsa-miR-624*, hsa-miR-501-3p and hsa-miR-101*, and downregulation of hsa-miR-183* in BPD-PH compared to sBPD samples **(Figure 4)**.

**Table 2.**
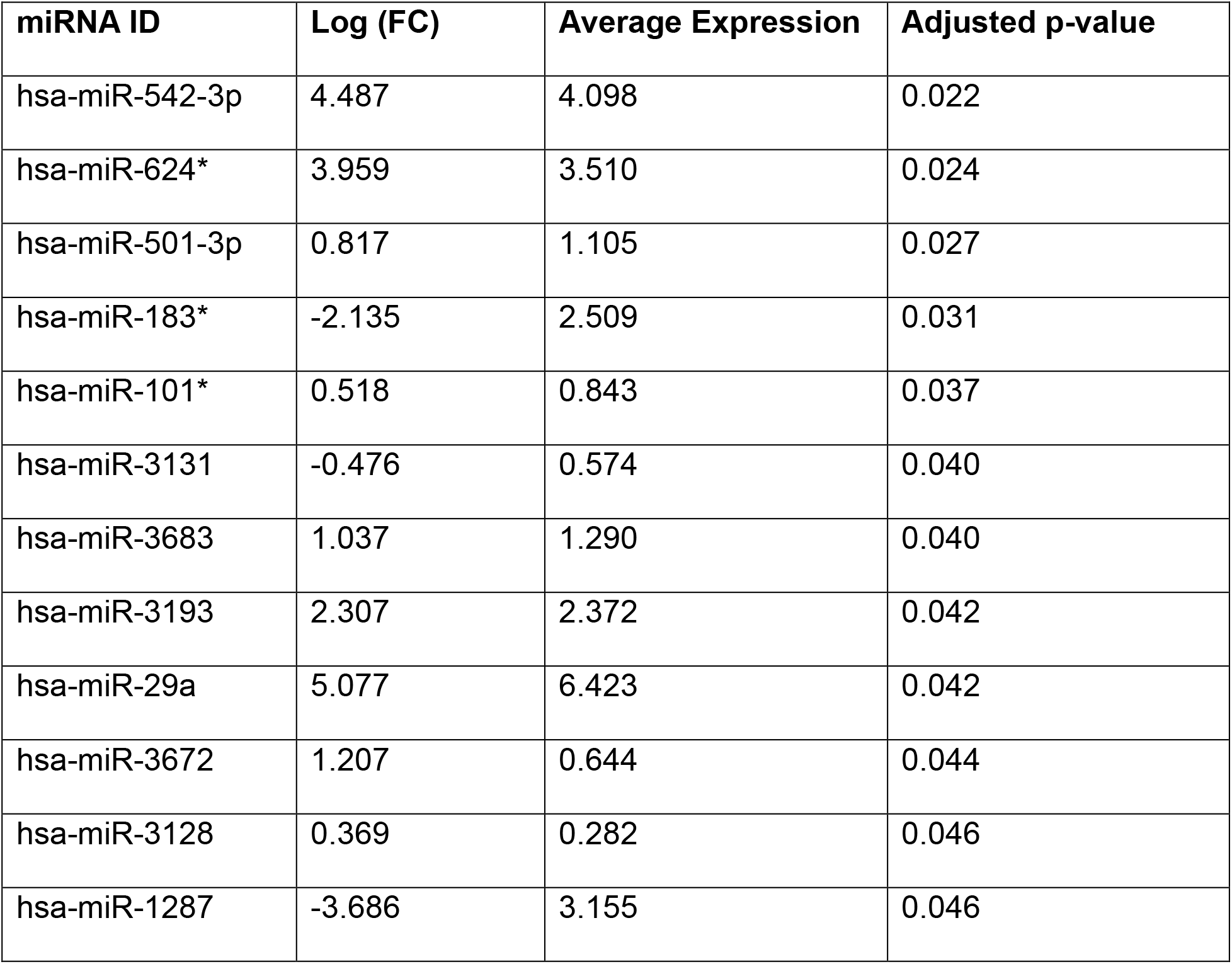
Differentially expressed miRNAs in BPD-PH vs. sBPD tracheal aspirates

| miRNA ID | Log (FC) | Average Expression | Adjusted p-value |
| --- | --- | --- | --- |
| hsa-miR-542-3p | 4.487 | 4.098 | 0.022 |
| hsa-miR-624* | 3.959 | 3.510 | 0.024 |
| hsa-miR-501-3p | 0.817 | 1.105 | 0.027 |
| hsa-miR-183* | -2.135 | 2.509 | 0.031 |
| hsa-miR-101* | 0.518 | 0.843 | 0.037 |
| hsa-miR-3131 | -0.476 | 0.574 | 0.040 |
| hsa-miR-3683 | 1.037 | 1.290 | 0.040 |
| hsa-miR-3193 | 2.307 | 2.372 | 0.042 |
| hsa-miR-29a | 5.077 | 6.423 | 0.042 |
| hsa-miR-3672 | 1.207 | 0.644 | 0.044 |
| hsa-miR-3128 | 0.369 | 0.282 | 0.046 |
| hsa-miR-1287 | -3.686 | 3.155 | 0.046 |

**Figure 1.**
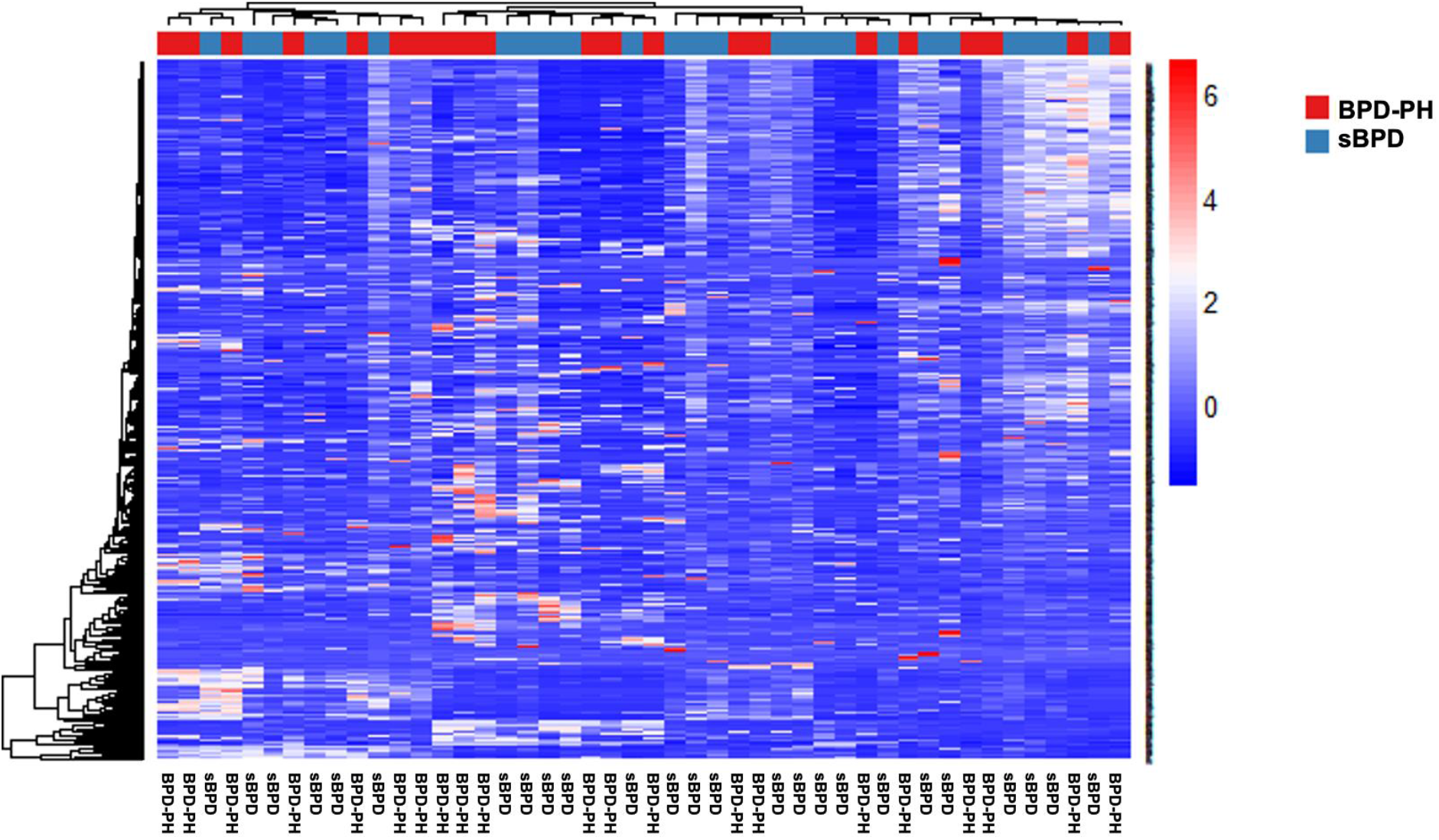
MiRNA expression in tracheal aspirates. Heatmap of expression of 1048 miRNAs in tracheal aspirates from infants with sBPD (n=25) and BPD-PH (n=21) (normalized by global mean), obtained by PCR arrays. Heatmap generated in R using the NMF package.

**Figure 2.**
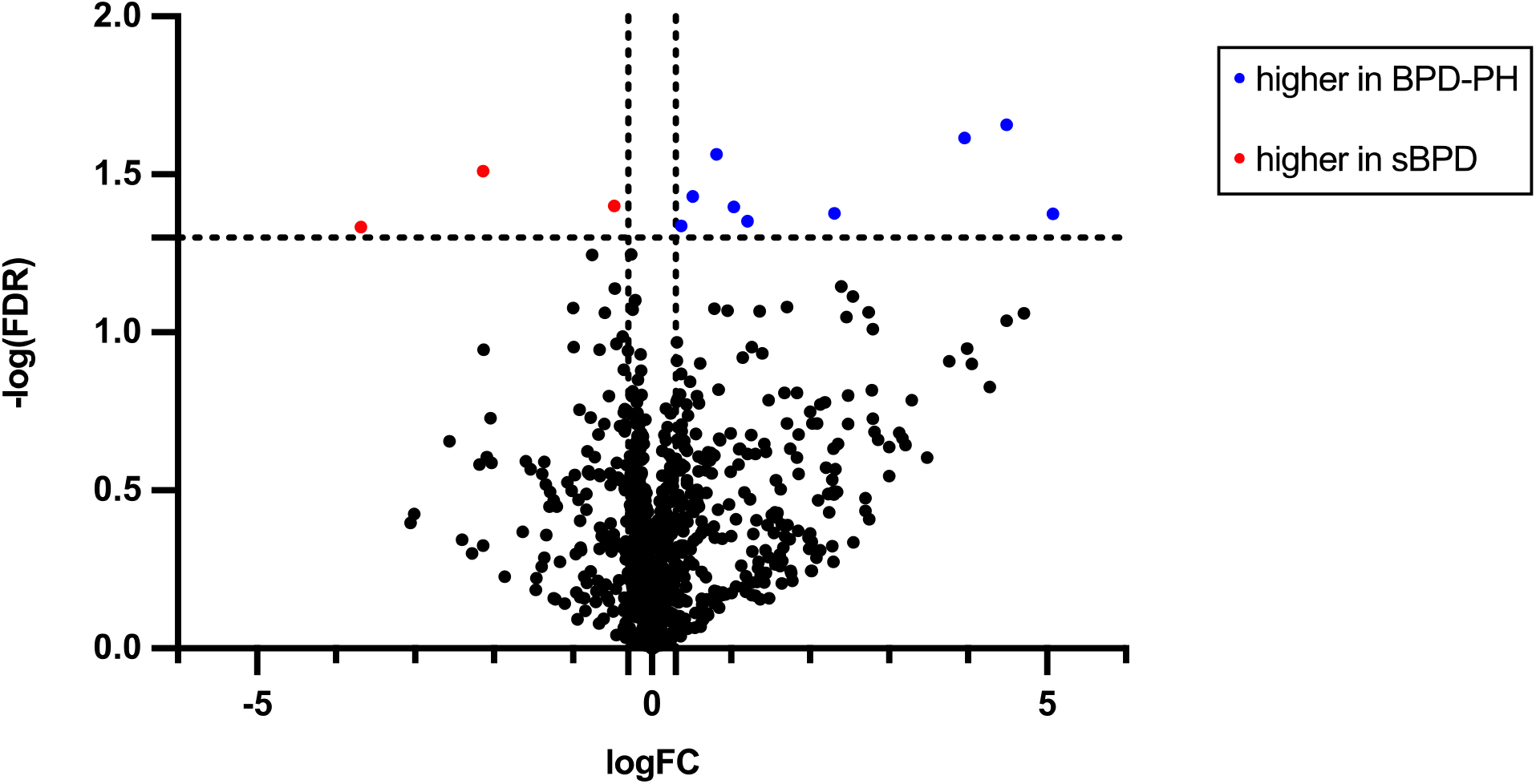
Volcano plot. Negative of false discovery rate vs. log of fold change of miRNAs between BPD-PH and sBPD groups. Vertical lines show |FC|=2 and horizontal line indicates FDR=0.05.

**Figure 3.**
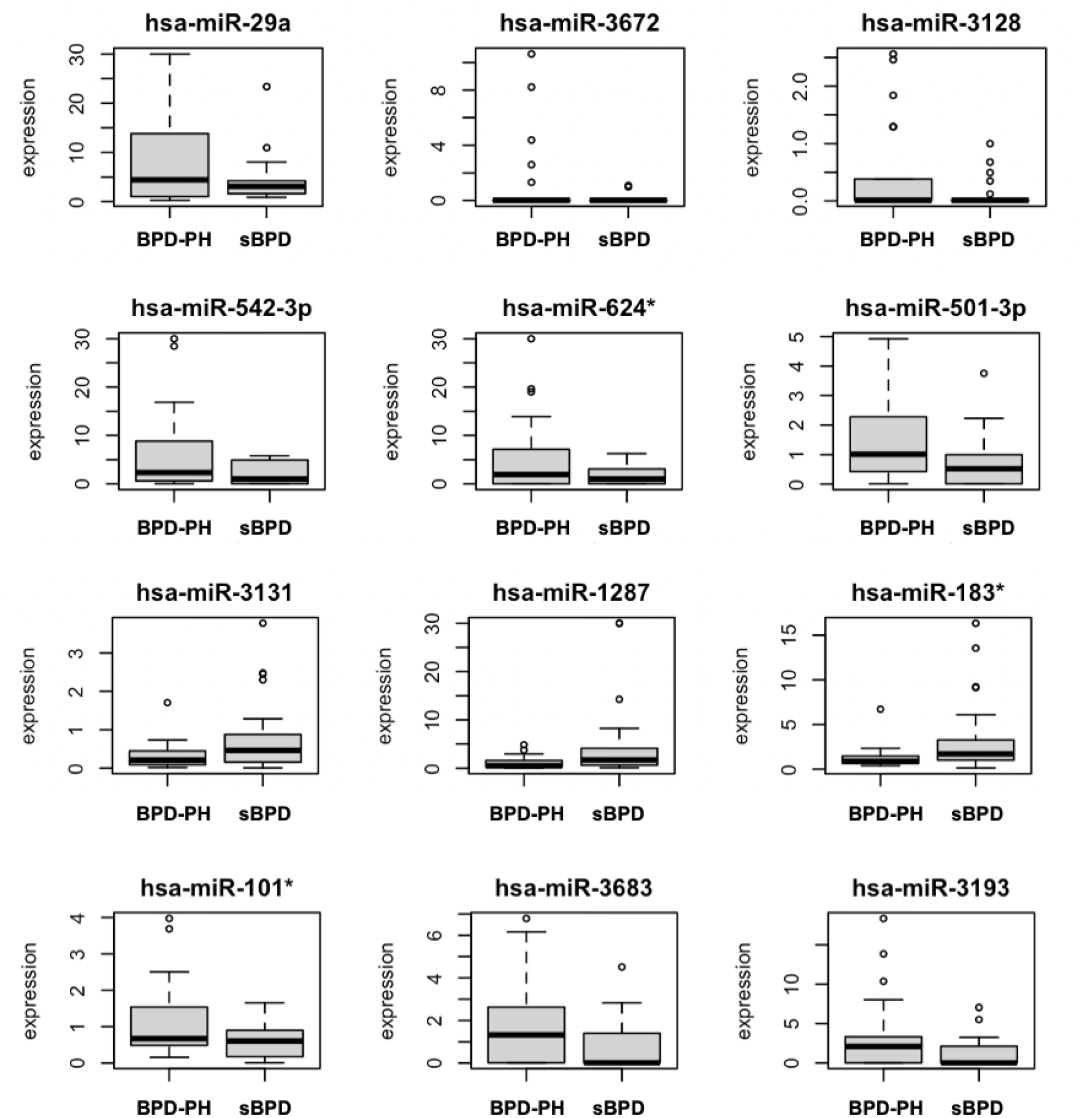
Top differentially expressed miRNAs. Y axes indicate relative expression values after global normalization. Graphs and calculations were done in R using the limma package

**Figure 4.**
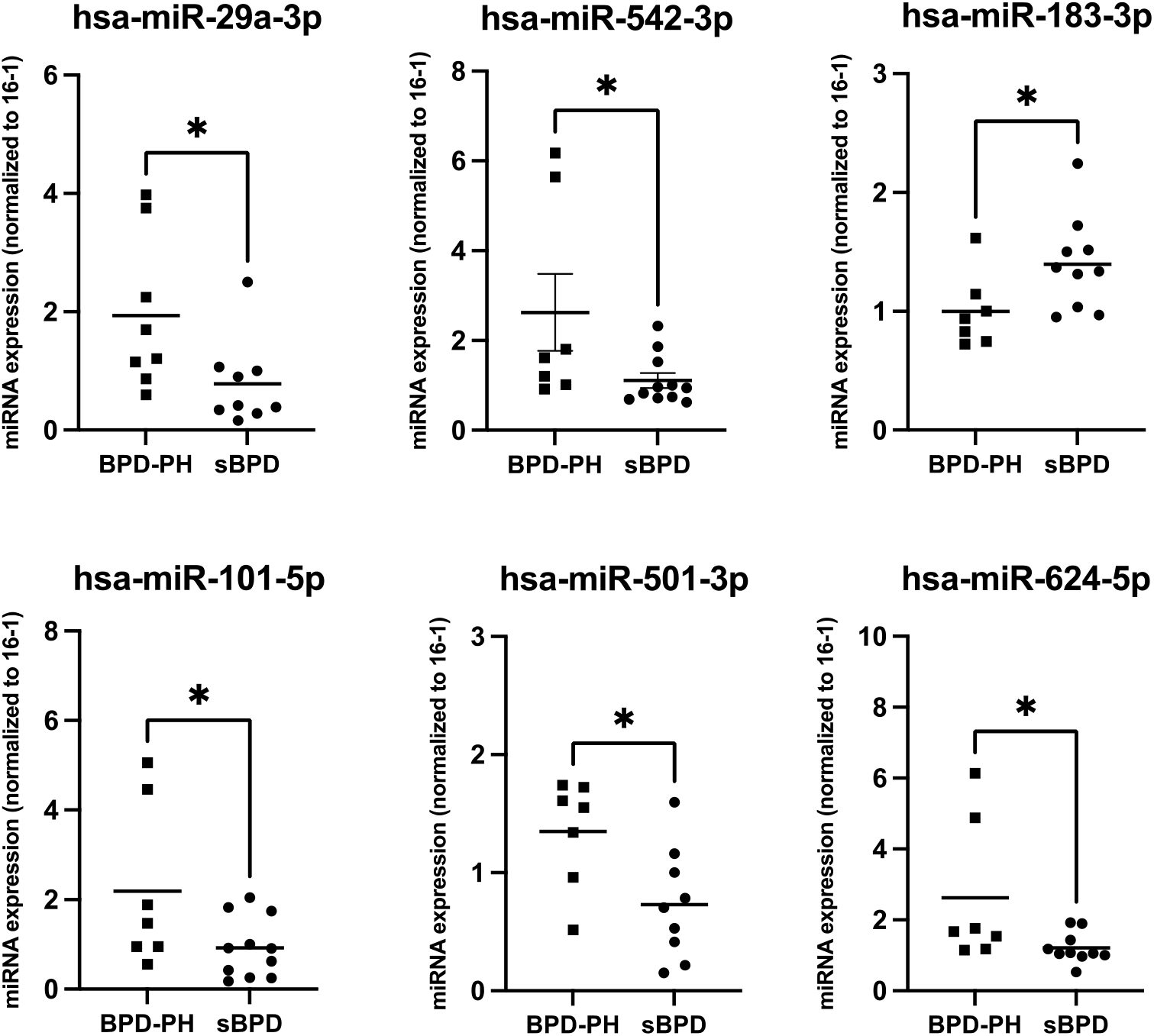
miRNA validation. Validation of miRNA expression by Real Time PCR. Y axes indicate relative expression values after normalization to miR-16, and a BPD-PH calibration sample using the 2^−ΔΔCT^ method (22, 23). Significant differences were determined by t-test using the GraphPad software (* p < 0.05)

### Transcriptomic analysis

The RNA seq analysis in 22 samples (7 BPD-PH and 15 sBPD) detected expression of 64,253 total transcripts in TAs. Of these, 31,420 transcripts showed counts >10. A heatmap of gene expression is shown in **Figure 5**, and a volcano plot is shown in **Figure 6**. A total of 1,584 genes displayed differential expression between groups (|logFC| > 2, p<0.05), including 221 with higher expression in BPD-PH vs sBPD and 1,003 with lower expression in BPD-PH vs. sBPD **(Supplementary Table 2)**. After correction for multiple comparisons, only 6 genes met the more stringent criteria for FDR<0.1 (IL6, RPL35P5, HSD3B7, RNA5SP215, OR2A1-AS1, and RNVU1-19) **(Table 3)**. Of these, IL6 had the highest differential expression and lower FDR, thus we validated its expression using qPCR in a subset of samples (**Figure 7**).

**Table 3.** Differentially expressed transcripts in tracheal aspirates of BPD-PH vs. sBPD patients

| Gene ID | Gene name | Log (FC) | p-value | FDR |
| --- | --- | --- | --- | --- |
| IL6 | Interleukin 6 | 4.372 | 4.777 | 5.55E-02 |
| RPL35P5 | Ribosomal Protein L35 Pseudogene 5 | 3.002 | 5.788 | 5.55E-02 |
| HSD3B7 | Hydroxy-Delta-5-Steroid<br>Dehydrogenase, 3 Beta- and Steroid<br>Delta-Isomerase 7 | 3.674 | 4.896 | 5.99E-02 |
| RNA5SP215 | RNA, 5S Ribosomal Pseudogene 215 | 3.362 | 5.230 | 5.99E-02 |
| OR2A1-AS1 | OR2A1 Antisense RNA 1 | -5.147 | 5.654 | 5.99E-02 |
| RNVU1-19 | RNA, Variant U1 Small Nuclear 19 | 3.397 | 5.062 | 5.99E-02 |

**Figure 5.**
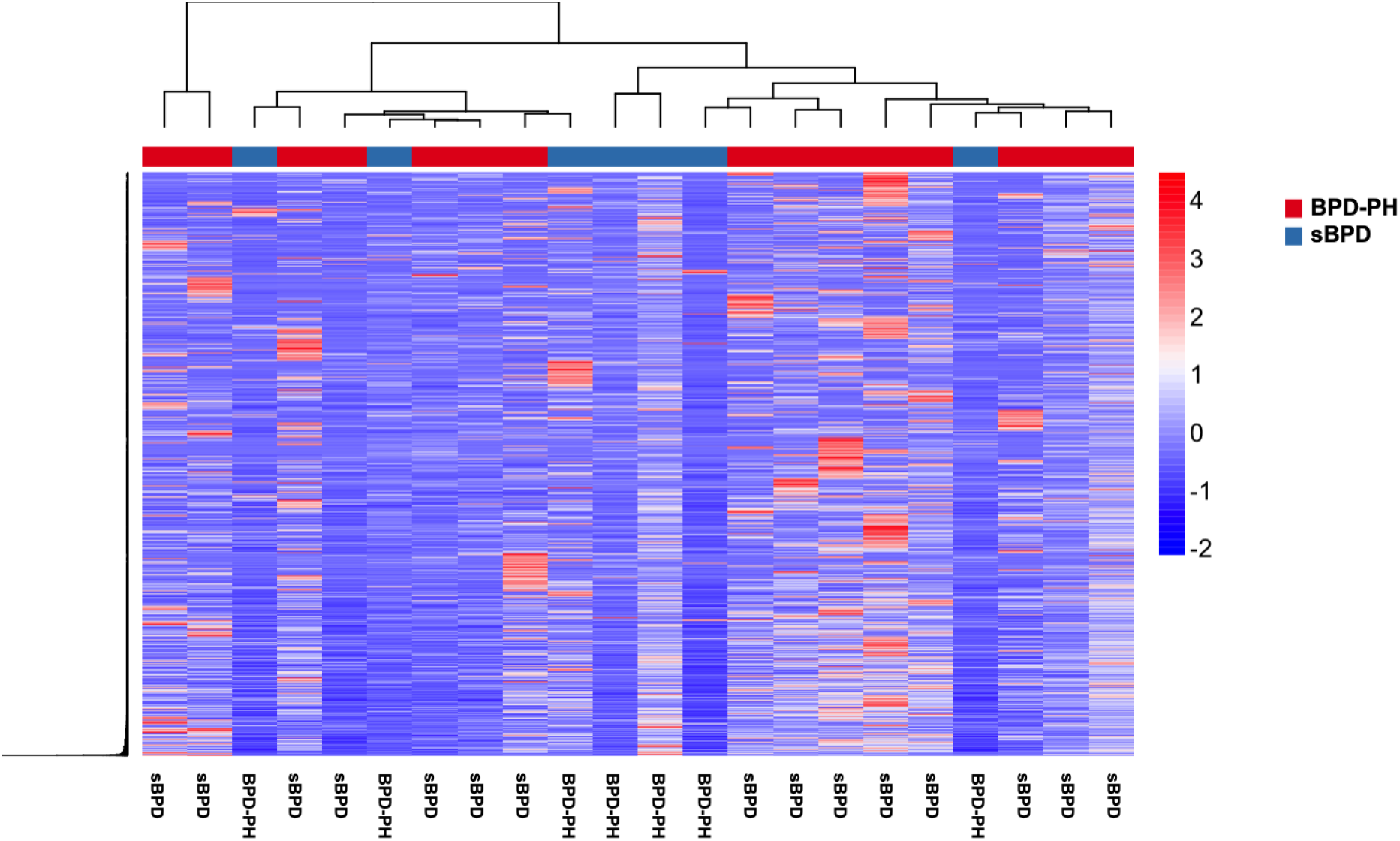
Transcriptomic analysis of tracheal aspirates. Heatmap of RNAseq counts (>10) for transcripts in TAs samples from BPD-PH (n=7) and sBPD (n=15) infants.

**Figure 6.**
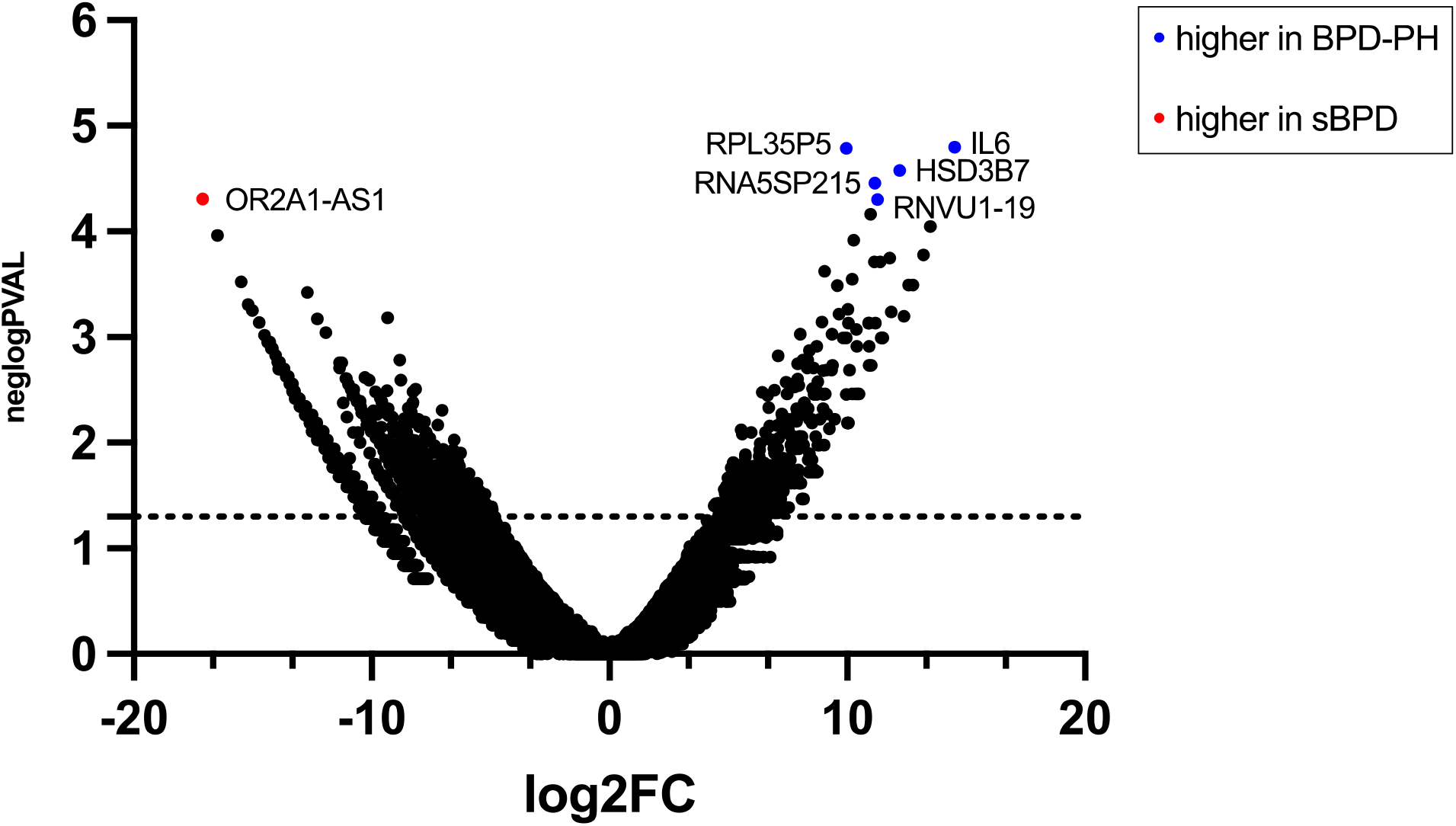
Volcano plot of mRNA transcripts. Negative log of false discovery rate vs. log of fold change of mRNAs between BPD-PH and sBPD groups. Horizontal line indicates FDR=0.05.

**Figure 7.**
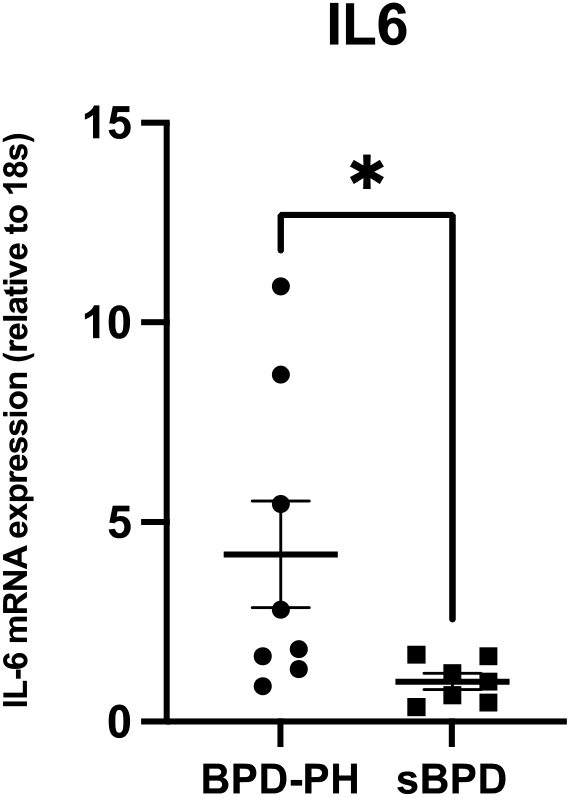
Interleukin-6 mRNA expression in tracheal aspirates. Validation of IL6 expression using Real-Time PCR shows increased expression in BPD-PH (n=8) samples compared to sBPD (n=7) (p<0.05)

### Proteomics analysis

Proteomics analysis was conducted in a subset of TA samples derived from 7 BPD-PH and 5 sBPD infants. A total of 712 different proteins were identified using the proteinpilot software. Of these, 64 were significantly different between groups (Mann Whitney Test p <0.05) (FDR<0.2). Eight proteins had at least 2-fold change in the BPD-PH group compared to the sBPD group **(Table 4)**.

**Table 4.** Differentially expressed proteins in tracheal aspirates of BPD-PH vs. sBPD patients

| **Proteins** | **Log (FC)** | **Mann-Whitney Test** | **Adjusted p-value** |
| --- | --- | --- | --- |
| calcyphosin isoform a | -2.24 | 0.021 | 0.041 |
| alpha-1-antitrypsin precursor | -2.26 | 0.005 | 0.055 |
| keratin, type II cytoskeletal 5 | -2.42 | 0.034 | 0.094 |
| pulmonary surfactant-associated protein B precursor | -3.11 | 0.009 | 0.110 |
| galectin-3-binding protein precursor | -4.04 | 0.020 | 0.114 |

### Pathway analysis

IPA analysis of differentially expressed miRNAs in BPD-PH vs. sBPD revealed significant associations with cell-to-cell signaling and interaction, cellular assembly and organization and cellular function and maintenance. The top physiological pathways associated with nervous system development and function, tissue development and cardiovascular system development and function. The top diseases and disorders associated were: cancer, organismal injury and abnormalities and gastrointestinal disease **(Table 5)**. IPA analysis of differentially expressed transcripts showed involvement of cellular development, cellular growth and proliferation and cell death and survival. The top physiological pathways were associated with embryonic development, hematological system development and function and hematopoiesis, and the top disease and disorders associated were cardiovascular disease, developmental disorders, and hereditary disorder **(Table 6)**. Similarly, IPA analysis of differentially expressed proteins identified molecular and cellular functions involving cellular compromise, protein synthesis, and cell death and survival. The top physiological pathway associated with identified proteins was tissue morphology, visual system development and function and organ morphology. The top diseases and disorders associated were inflammatory response, cancer and organismal injury and abnormalities **(Table 7)**. Finally, the integrated pathway analysis of differentially expressed miRNAs, mRNAs and proteins highlighted NFkB complex, VEGF, SERPINA1, as well as insulin, LDL, ERK1/2 and IL6 as the affected pathways in BPD-PH when compared to sBPD **(Figure 8**).

**Table 5:** IPA pathway associated with differentially expressed miRNA in BPD-PH versus sBPD

| **Molecular and Cellular Functions** | **P-Value Range** | **Number of Molecules** |
| --- | --- | --- |
| Cell-To-cell Signaling and Interaction | 1.27E-03 – 5.08E-04 | 2 |
| Cellular Assembly and Organization | 2.03E-03 – 5.08E-04 | 2 |
| Cellular Function and Maintenance | 1.54E-02 – 5.08E-04 | 2 |
| Cellular Development | 4.95E-02 – 8.44E-04 | 4 |
| Cellular Growth and Proliferation | 4.95E-02 – 8.44E-04 | 4 |
| **Physiological System Development and Function** |  |  |
| Nervous System Development and Function | 8.87E-03 – 5.08E-04 | 2 |
| Tissue Development | 1.54E-03 – 5.08E-04 | 3 |
| Cardiovascular System Development and Function | 1.27E-03 – 1.27E-03 | 1 |
| Connective Tissue Development and Function | 4.56E-03 – 1.27E-03 | 2 |
| Hematological System Development and Function | 1.54E-03 – 1.27E-03 | 1 |
| **Diseases and disorders** |  |  |
| Cancer | 4.47E-02 – 1.56E-04 | 5 |
| Organismal Injury and Abnormalities | 4.47E-02 – 1.56E-04 | 6 |
| Gastrointestinal Disease | 4.20E-02 – 3.19E-04 | 5 |
| Respiratory Disease | 3.19E-04 – 3.19E-04 | 2 |
| Tumor Morphology | 2.54E-03 – 1.02E-03 | 1 |

**Table 6:**
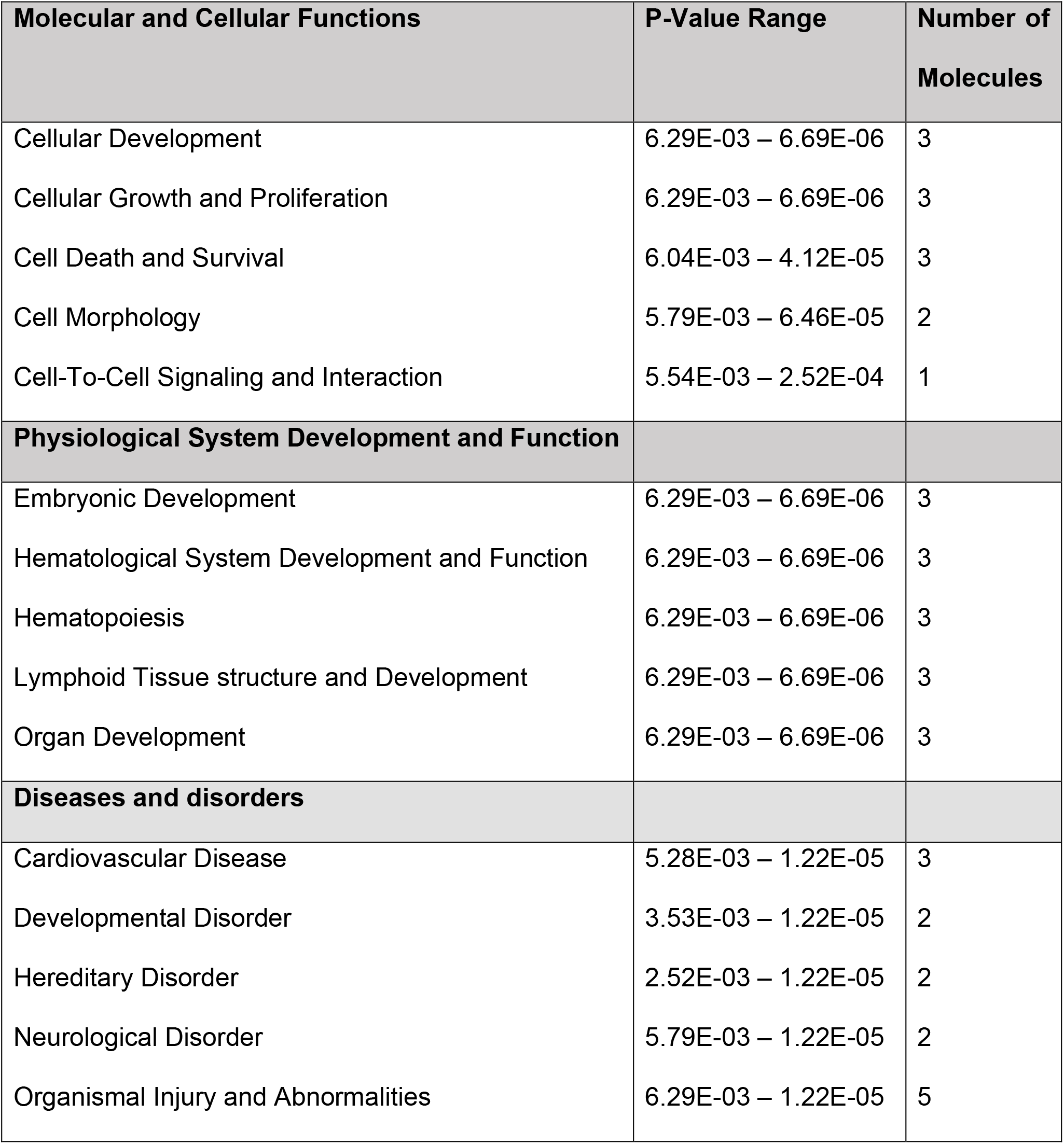
IPA pathway associated with differentially expressed mRNA in BPD-PH versus sBPD

**Table 7:** IPA pathway associated with differentially expressed proteomics in BPD-PH versus sBPD

| **Molecular and Cellular Functions** | **P-Value Range** | **Number of Molecules** |
| --- | --- | --- |
| Cellular Compromise | 2.41E-02 – 5.94E-10 | 8 |
| Protein Synthesis | 2.18E-02 – 2.71E-06 | 7 |
| Cell death and Survival | 2.10E-02 – 1.82E-05 | 9 |
| Cell Morphology | 1.63E-02 – 2.70E-05 | 8 |
| Cellular Movement | 2.32E-03 – 2.70E-05 | 7 |
| **Physiological System Development and Function** |  |  |
| Tissue Morphology | 1.76E-02 – 6.03E-06 | 6 |
| Visual System Development and Function | 6.04E-07 – 6.04E-07 | 3 |
| Organ Morphology | 2.36E-02 – 9.02E-06 | 5 |
| Organismal Development | 2.36E-02 – 9.02E-06 | 7 |
| Reproductive System Development and Function | 2.04E-02 – 9.02E-06 | 2 |
| **Diseases and disorders** |  |  |
| Inflammatory Response | 2.32E-02 – 5.94E-10 | 10 |
| Cancer | 2.29E-02 – 6.03E-07 | 12 |
| Organismal Injury and Abnormalities | 2.45E-02 – 6.03E-07 | 12 |
| Reproductive System Disease | 2.43E-02 – 6.03E-07 | 8 |
| Developmental Disorder | 1.99E-02 – 7.85E-06 | 9 |

**Figure 8.**
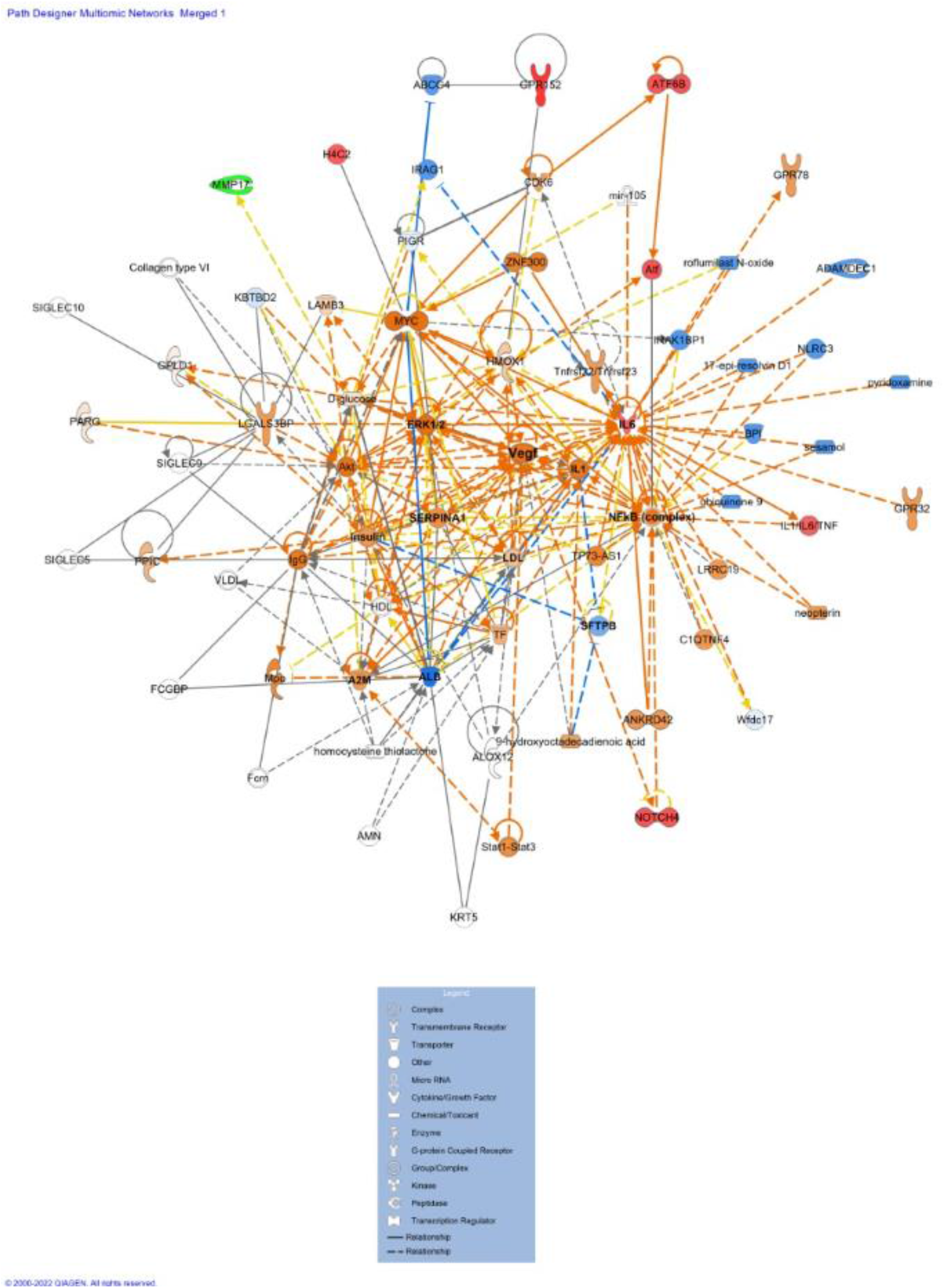
IPA pathways. Ingenuity pathway analysis of the differentially expressed miRNAs, proteins, and mRNAs in TAs of sBPD and BPD-PH

## Discussion

Pulmonary hypertension associated with bronchopulmonary dysplasia is a multifactorial disease with high morbidity and mortality (36). The mechanisms that differentiate the pathophysiology of sBPD and BPD-PH development remain unknown, which limits our ability to properly diagnose and treat infants (37). Using a multi-omic approach, our study has identified a novel biomarker panel including 12 miRNAs, 6 transcripts, and 5 proteins which distinguish infants with BPD-PH from those with sBPD who lack detectable PH on echocardiogram. Target molecules identified via IPA are NFkB, VEGF, SERPINA1, IL6, ERK12 and Insulin to name a few. Careful analysis of relevant individual markers identified a recurring theme of angiogenesis, hyperoxic stress and vascular remodeling as some of the key pathways that help endotype infants with severe lung disease with and without PH.

Identifying disease endotypes using biomarkers will help not only identify underlying mechanistic pathways, but also help clinically stratify high risk infants to help improve long term outcomes. For this approach to have translational value, it is important to identify an affordable, minimally invasive, bedside point of care biomarker. Our study using TA samples in preterm infants leverages a noninvasive easily obtained biofluids. This could be a valuable tool to stratify at-risk infants and help clinical decision making.

Multi-omics approaches have been used to understand complex pulmonary disease such as asthma, COPD, ARDS, IPF and PAH in order to develop personalized diagnostics and treatments (38). While some studies have used Arginine metabolites to endotype pulmonary arterial hypertension (39), we chose to conduct a non-targeted approach, given that there is a complex array of prenatal and postnatal stressors associated with BPD-PH that can result in arrested pulmonary vascular development. While antenatal and perinatal factors including intrauterine growth restriction, preeclampsia, maternal chorioamnionitis, and hypoxic and hyperoxic stress responses lead to abnormal pulmonary vasculogenesis and alveolarization that have been associated with sBPD, BPD-PH presents with vascular disease in its most severe form (40, 41). This suggests that BPD-PH is a more severe spectrum of sBPD phenotype.

Our analysis revealed 9 miRNAs that were upregulated in BPD-PH when compared to sBPD. Of particular interest, miRNA hsa-miR-29a, which is upregulated in BPD-PH, has been previously linked with vascular disease. A publication by Chen et. al showed upregulation of miRNA 29 by 16α-Hydroxyestrone resulting in BMPR2 associated pulmonary hypertension both in human and animal models (42). Moreover, miRNA-29a is also a well-studied biomarker for hypertrophic cardiomyopathy (43), and thus could indicate myocardial stress as a result of elevated pulmonary pressure. Another study has noted microvesicle-secreted miR-29a/c significantly suppresses VEGF expression in gastric cancer, inhibiting vascular cell growth (44). Contradicting, in a pre-eclampsia model of human umbilical vein endothelial cells, knockdown of miRNA 29a/c-3p inhibited VEGF2 and FGF2 induced endothelial migration (45). Bhatt and colleagues demonstrated reduced expression of VEGF mRNA along with VEGF receptor in preterm infants with fatal lung disease (46), while Lassus and colleagues demonstrated lower VEGF in preterm infants with persistent severe lung disease when compared to those who recovered (16). Whether miRNA 29a is a biomarker of VEGF expression or cardiac stress in preterm infants with BPD-PH needs further exploration.

Another miRNA highly expressed in BPD-PH vs. sBPD was hsa-miR-542-3p. This miRNA has been shown to target BMP7 and regulate osteogenic transition of vascular smooth muscle cells in aging and hepatic fibrosis in rats (47, 48). Additionally, of the 3 down regulated miRNAs, hsa-miR-183 is typically expressed as a cluster miR-183-96-82 and packaged in exosomes expressed in high levels in cancer types including breast and prostate (49). This miRNA functions as an oncogene by targeting transcription factor EGR1 and promote tumor migration (50) and potentially inhibits NFKB-1 expression by directly targeting its 3’-untranslated region (51). Similarly, hsa-miR-101-5p has been studied to regulate cell proliferation through inhibition of RAP1A, which is involved in embryonic blood vessel formation (52). This miRNA has also been shown to inhibit growth, proliferation, and migration through targeting Sex-determining Region Y-Box 2 (SOX2) (53). Overall, our miRNA analysis revealed multiple potential key players of the mechanisms involved in BPH-PH progression.

While our transcriptomic analysis identified multiple pseudogenes, and non-coding RNAs as the top differentially expressed transcripts between BPD-PH and sBPD, it also revealed IL6 as a highly expressed gene in TAs from infants with BPD-PH. Previously, a multicenter study identified IL6 as a clinical marker to predict severe phenotype in pediatric pulmonary arterial hypertension (54). This cytokine has been found to modulate expression of BMPR2 (55) similar to miRNA 29a which plays a role in vascular remodeling (56). In addition, IL6 is shown to be an important clinical marker for clinical phenotype and survival in patients with pulmonary hypertension. Another differentially expressed gene identified was HSD3B7 which has been shown to regulate endometrial activity in women with polycystic ovarian syndrome (57) who’s novel mutation is found to be associated with neonatal cholestasis (58).

Finally, our proteomic analysis revealed Galectin 3 binding protein precursor gene as the most differentially expressed protein. Galectin is an important mediator of VEGF and bFGF mediated angiogenic response which has been shown to be reduced in the presence of Galectin 3 inhibitors (59, 60). Another protein of interest was Surfactant protein B, as gene polymorphisms of SFPB have shown to be associated with BPD and newborn respiratory distress syndrome (61, 62).

The IPA analysis conducted separately for the differentially expressed miRNAs, mRNAs and proteins showed cellular movement, cellular development and cell-to cell signaling as the common top molecular functions suggesting pathways regulating growth and organ development. The analysis revealed that the top physiological systems were nervous system development and function, embryonic development and tissue morphology.

Some of the strengths of our study are our non-biased approach to identify biomarkers to endotype preterm infants with sBPD vs. BPD-PH. Using a multi-omic approach helped us identify coding and non-coding, as well as transcriptional, and post transcriptional markers. We used bioinformatic tools to identify key pathways and mechanisms converging and comparing the miRNAs, mRNA and proteomic data. Given the complex pathophysiology of sBPD and BPD-PH, our study combining biological fluids with a multi-omic approach identified predicted targets that have been culminated by various pathways resulting in sBPD and BPD-PH phenotypes. To ensure inter reporting bias, all echocardiograms were reviewed by a single pediatric cardiologist. We excluded ELGANs with hemodynamically significant intracardiac shunt in the study to eliminate confounding factors resulting from pulmonary over circulation or sheer stress on pulmonary vasculature.

Despite our findings, this study is not without limitations. As a pilot study, the size of the study cohort leaves opportunity for a larger follow up study. However, while a study size of 46 preterm infants seems like a smaller cohort, it is very challenging to recruit and conduct sample analysis from ELGANs especially those who are invasively ventilated, since the clinical preference is to noninvasively ventilate which limits our access to tracheal aspirate samples. Echocardiograms were used as the diagnostic tool, which could be criticized as it lacks the sensitivity and specificity of the gold standard cardiac catheterization; however, we point out the limitation of reliance on invasive cardiac catheterization in extreme preterm infants which is a population less likely to undergo cardiac catheterization. There have been multiple studies validating specific echocardiogram parameters with high correlation with cardiac catheterization (63-65). Identifying pathological biomarkers would help increase the sensitivity and specificity of non-invasive tools such as echocardiograms which measure indirect evidence of pulmonary vascular resistance.

In conclusion, there is a significant overlap in the pathophysiology of sBPD and BPD-PH along with their clinical signs and symptoms. Our study evaluated and identified specific multi-omic markers in tracheal aspirate of ELGANS that aid in specific endotyping of the complex disease of pulmonary vascular disease associated with BPD in these infants. We report specific miRNAs, transcripts, and proteins that could potentially serve as markers of angiogenesis and inflammatory biomarkers of pulmonary vascular disease pathology in preterm infants with BPD.

## Data Availability

All data produced in the present work are contained in the manuscript as links to online repository

https://www.ncbi.nlm.nih.gov/geo/query/acc.cgi?acc=GSE205138

## Acknowledgements

The authors would like to thank the Penn State Health Proteomics Core facility staff, and its director Dr. Bruce Stanley for proteomic analysis. We also thank Diane Kitch for data coordination and IRB approval assistance, as well as Debra Spear and Susan DiAngelo for sample collection and processing, and Mathew Steadman and Dr. Chongben Zhang from the University of North Carolina at Chapel Hill Biobehavioral Core facility for sample storage and processing.

## Notes

### Competing Interest Statement

The authors have declared no competing interest.

### Funding Statement

Center for Research on Women and Newborn Health; Childrens Miracle Network; Department of Pediatrics Startup funding; Penn State Health Core facility laboratory

### Author Declarations

Penn State Health institutional review board (STUDY 00000482) gave ethical approval for this work

